# Changes in presentation, treatment, and survival of patients with hepatocellular carcinoma in Damietta, Egypt, 2007-2019: a retrospective monocentric cohort study

**DOI:** 10.1101/2022.09.07.22279666

**Authors:** Kévin Jean, Ahmed Tawheed, Liem Binh Luong Nguye, Tarek Heikal, Usama Eldaly, Neven Elhadidy, Ahmed Elghaieb, Ahmed Aboudonia, Laura Tondeur, Amélie Dublineau, Arnaud Fontanet, Mohamed El-Kassas

## Abstract

**Context:** We aimed to assess temporal changes in the presentation and survival of patients with hepatocellular carcinoma (HCC) in the northern Egypt region, one of the regions reporting the highest incidence of the disease globally.

**Method:** We conducted a monocentric retrospective study. Patients presenting at the Damietta Oncology referral center between 2007 and 2019 with a diagnosed HCC were eligible. Individual, clinical and tumor characteristics at HCC diagnosis, including the Barcelona Clinic Liver Cancer (BCLC) staging, were retrieved from medical files and patients’ final vital status was ascertained by combining various data sources. Patients were divided into 2 groups based on diagnosis period: pre- and post-2014. Survival was analysed based on Kaplan-Meier curves and differences in restricted mean survival time (RMST).

**Results:** Data from 5,097 patients (among 5,210 eligible, 97.8%) wereere analyzed. We observed a significant trend toward HCC diagnosed at earlier stage in the post- vs. pre-2014 period (BCLC stage 0/A or B: 37.2% vs. 27.1%, p<10^−3^). Overall patient’s survival after the HCC diagnosis was poor, with a median of 8.1 months. The BCLC staging system performed well in predicting survival (median survival for patients diagnosed with BCLC stage 0/A, B, C, and D: 28, 25, 7, and 4 months, respectively, p<10^−3^). Despite a trend toward HCC diagnosed at earlier stages, we did not observe a significant improvement in survival over time. Overall, treatments offered in this medical center were in line with international guidelines, and 16.1% of the patients who received a curative treatment had an improved survival (30.7 months in median). However, HCC recurrence was frequent among patients cured for HCC, with a median time to recurrence of 22 months.

**Conclusion:** Overall survival after HCC diagnosis in Egypt remains poor but is significantly improved by curative therapy. Despite a trend toward earlier diagnosis of HCC, we did not observe a general improvement in survival over time, which remains to be clearly understood.

---

Hepatocellular carcinoma (HCC) is the fifth most common cancer globally but occupies the first place among cancers in Egypt [1]. This special situation is primarily explained by the unique scale of the Hepatitis C Virus (HCV) epidemic in the country [2], which has been historically associated with the mass parenteral anti-schistosomal treatment campaigns in the 1960s–70s [3] and more recently with unsafe medical practices [4]. Moreover, the incidence rates for HCC in Egypt show geographical variation matching HCV prevalence, with the highest figures in the northern Delta region compared to the lowest in the southern region [1]. Untreated chronic HCV infection may progress into cirrhosis and liver cancer, for which the prognostic is very poor.

Previous studies documented significant disparities in the survival of patients with HCC according to individual characteristics, such as age, gender, ethnicity, and socioeconomic status [5–7]. HCC etiology may also affect response to treatment and survival [8]. Other conditions, such as metabolic disorders, may affect HCC incidence, but their influence on survival is not clearly established [9]. Better identification of individual prognostic factors among patients with HCC in the specific context of Egypt may provide valuable insight into patients’ care and treatment.

Furthermore, recent advances in HCC and HCV treatment may have changed the overall survival of patients with HCC. Tumor staging at the time of diagnosis is critical to estimate patient prognosis and choose between treatment options. Several staging systems for HCC have been developed [10], among which the Barcelona Clinic Liver Cancer (BCLC) has been endorsed by the European Association of the Study of the Liver (EASL) and the American Association of the Study of Liver Diseases (AASLD) [11]. The BCLC staging system has been documented to perform well within a monocentric Egyptian study [12]. Confirming these results in the healthcare center acknowledging the highest incidence of HCC in Egypt may be valuable for the national treatment guidelines [13].

Moreover, the availability of direct-acting antivirals (DAA) through the Egyptian National Treatment Programs has encouraged HCV testing actions (screening for more than 65 million population over 8 months) and the linked HCC screening activities among those infected with HCV, especially cirrhotic patients, over the past years [14,15]. This is expected to have led to earlier HCC diagnosis and better prognostic overall. Additionally, the large number of treated HCV patients in Egypt (more than 4 million) may impact the landscape of HCC in the country. Documenting such trends –if they exist– will encourage further efforts and help assess the remaining benefits that may be expected from expanded HCV testing.

In this monocentric retrospective study, we assessed temporal changes in the presentation and survival of patients presenting with HCC between 1^st^ Jan 2007 and 31^st^ Dec 2019 in a tertiary specialized oncology referral center in Damietta, northern Egypt.

## METHODS

The Damietta Oncology Center (DOC) is one of the major tertiary specialized oncology referral centers in northern Egypt and serves five large governorates in the Delta region. The Center covers the region with the highest incidence of HCC in Egypt [13]. In 2013, Damietta Oncology Center launched a multidisciplinary HCC unit outfitted with necessary equipment for the planned interventions. A specialized HCC multidisciplinary clinic is responsible for consultations for HCC cases with hepatology, radiology, and surgical oncology consultants. All patients were recruited and diagnosed according to the BCLC guidelines [16]. Baseline demographic, clinical, laboratory and radiological criteria were documented in the patient file. Additionally, the BCLC staging was calculated and documented. Finally, a patient management plan is decided according to the BCLC staging and considering the logistics and availability of various treatment options in the center.

### Recruitment

All patients presenting between 1^st^ Jan 2007 and 31^st^ Dec 2019 at the DOC with a diagnosis of HCC were eligible for the study.

Four trained medical doctors from Damietta Oncology Centre were responsible for the active data collection from the archived medical records of the patients. The study’s data manager regularly revised the data collection process. The extracted data were transformed into a unique pre-prepared case report form designed for this study, which included general, tumor-related, imaging, and laboratory data. The case report forms were electronically submitted to a web-based software (REDCap) developed explicitly for online data entry, validity checks, and preparation for data analysis. Death registries in local health governorates were regularly checked for reported deaths of eligible patients. Follow-up phone calls were scheduled for patients who missed their follow-up or those with an unknown situation. Electronic checks were performed using Check Programs. Data entry was done depending on the Hospital Identification Number to prevent duplicates.

### HCC diagnosis

The date of HCC diagnosis was determined using non-invasive imaging criteria as described by international recommendations, and BCLC stage was ascertained upon diagnosis [17–19]. HCC etiology was categorized based on hepatologic criteria as HCV, HBV, HBV-HCV coinfection, or non-viral etiology. The presence of anti-HCV determined hepatitis C status; hepatitis B status was determined by the presence of hepatitis B surface antigens.

### Definition of HCC primary treatment

HCC primary treatment was defined as the most advanced level of treatment received, categorized herein in decreasing order: surgical liver resection, curative locoregional therapy (radiofrequency or microwave ablation, percutaneous ethanol injection), noncurative locoregional therapy (transarterial chemoembolization), and supportive care only.

### Final status

The final vital status and date of last contact or death were ascertained following three steps. First, the medical file was searched for information regarding the date of last contact and notification of death. Patients with date of latest notice <3 months at the date of data collection were considered as alive. For patients with date of latest notice ≥3 months, the Egyptian national Cancer registry was consulted for patients’ information, and if occurring, the date of death was retrieved. Third, for patients with no explicit mention of death as per the medical file or cancer registry, the research team tried to contact the patient or patient’s next of kin based on contact data in the medical file. The research team attempted contact up to three times at different dates and times to collect information regarding patients’ vital status and date of death, if any. Patients with unsuccessful contact procedures were finally considered alive at the latest date appearing in the medical file.

### HCC recurrence-free survival after curative treatment

We assessed the HCC recurrence-free survival among patients who received curative therapy (either surgical liver resection or curative locoregional therapy). Among these, we defined recurrence-free survival as starting at the date of treatment end ending at the recorded date of recurrence or being censored.

### Statistical analysis

All eligible patients were included in the statistical analysis provided they had a non-missing date of diagnosis and date of point. Descriptive statistics were reported as percentage for categorical variables and as median with 25^th^ and 75^th^ percentiles for continuous variables. We used the Chi-2 test to test for changes in the distribution of categorical variables across time period of HCC diagnosis and the Student test for continuous variables. The Kaplan-Meier method was used to plot survival curves, and the log-rank test was used to assess survival difference in univariate analysis. Classical survival analysis based on Cox regression estimates hazard ratio, which is difficult to interpret when the proportional hazards assumption is violated. Moreover, the shortcomings of the hazard ratio are being increasingly recognized [20,21]. We therefor analysed the association between individual characteristics and all-cause mortality using difference in restricted mean survival time (RMST) with 95% confidence intervals (CI) using the *survRM2* R package [22]. In the multivariate analysis, we only included variables that can be assessed upon admission and that are not considered within the BCLC staging system. Presentation with HCC and survival was analysed overall and also while stratifying on the date of HCC diagnosis. The period of HCC diagnosis was categorized into before/after 2014, which corresponds

In the multivariable analysis, we only included variables that can be assessed upon admission and that are not considered within the BCLC staging system. Presentation with HCC and survival was analysed overall and also while stratifying on the date of HCC diagnosis. The period of HCC diagnosis was categorized into before/after 2014, corresponding to the initiation of the national DAA scale-up program [14].

## RESULTS

Medical records were retrieved among 5,210 eligible patients having presented at the DOC between 1^st^ Jan 2007 and 31^st^ Dec 2019, for whom data was collected and vital status ascertained (Supplementary Figure 1). Among these, 5,097 (97.8%) had a non-missing date of diagnosis and date of latest notice and were included in the statistical analysis. The number of patients diagnosed was 178 in 2007 (with one diagnosed in 2005 and 6 diagnosed in 2006), peaked at 654 in 2016 and decreased at 550 in 2018 (Figure 1).

**Figure 1:**
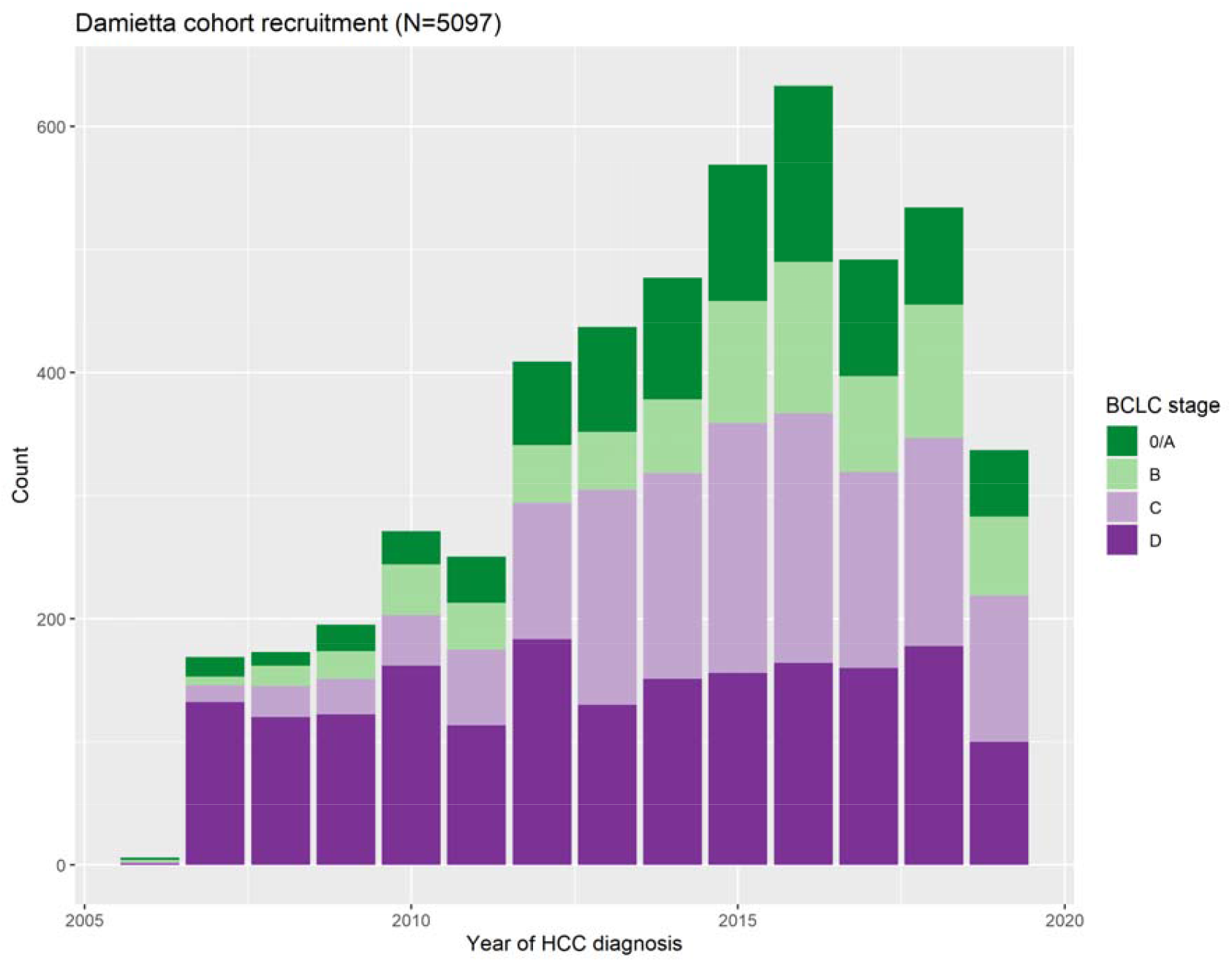
Distribution of BCLC stage at HCC diagnosis, by year of diagnosis.

### Temporal changes in presentation and treatment option

Table 1 presents patients’ general and clinical characteristics, overall and by period of HCC diagnosis. Overall, 5,165 (99.3%) patients were cirrhotic, and only 3 patients (<0.1%) reported any alcohol consumption. Over time, the proportion of patients from urban area with hypertension or diabetes increased; the proportion of patients reporting tobacco consumption or with ascites decreased. The proportion of patients of ECOG performance status from 0 to 2 increased over time (Supplementary Figure 2). Overall, the etiology of HCC was linked to HCV in 91.6% of the patients, linked to HBV in 3.3%, linked to HBV-HCV coinfection in 0.8%, and non-viral in 4.3%, with no significant changes over time (p=0.22).The large majority were diagnosed using CT scan. The number of hepatic focal lesions did not change significantly over time. The size of the largest HFL tended to decrease, but this variable was affected by a high proportion of missing values. The proportion of patients with distant metastasis tended to decrease over time (p=0.092).

**Table 1 :** Temporal changes in patients’ general, clinical, and tumor characteristics at HCC diagnosis.

|  |  | Year of HCC diagnosis |  |  |  |
| --- | --- | --- | --- | --- | --- |
|  |  | Overall | 2005-2013 | 2014-2019 |  |
|  |  | N = 5,097 | N = 2,465 | N = 2,632 | P-value |
| General characteristics |  |  |  |  |  |
| Sex | Male | 3947 (77.4%) | 1929 (78.3%) | 2018 (76.7%) | 0.1764 |
|  | Female | 1150 (22.6%) | 536 (21.7%) | 614 (23.3%) |  |
| Area of residence | Urban | 1284 (25.2%) | 544 (22.1%) | 740 (28.1%) | <10<sup>-3</sup> |
|  | Rural | 3809 (74.8%) | 1920 (77.9%) | 1889 (71.9%) |  |
|  | NA | 4 | 1 | 3 |  |
| Age at 1st visit in DOC | Median [Q25-Q75] | 61 [55-67] | 59 [53-66] | 62 [56-67] | <10<sup>-3</sup> |
|  | NA | 4 | 2 | 2 |  |
| Tobacco consumption | No | 3316 (65.2%) | 1486 (60.4%) | 1830 (69.7%) | <10<sup>-3</sup> |
|  | Yes | 1769 (34.8%) | 973 (39.6%) | 796 (30.3%) |  |
|  | NA | 12 | 6 | 6 |  |
| Clinical characteristics |  |  |  |  |  |
| Hypertension | No | 3850 (76.4%) | 1970 (81.2%) | 1880 (72.1%) | <10<sup>-3</sup> |
|  | Yes | 1186 (23.6%) | 457 (18.8%) | 729 (27.9%) |  |
|  | NA | 61 | 38 | 23 |  |
| Diabetes | No | 3721 (73.9%) | 1882 (77.6%) | 1839 (70.4%) | <10<sup>-3</sup> |
|  | Yes | 1316 (26.1%) | 544 (22.4%) | 772 (29.6%) |  |
|  | NA | 60 | 39 | 21 |  |
| History of gastroesophageal varices | No | 3937 (83.0%) | 1952 (83.2%) | 1985 (82.8%) | 0.6939 |
|  | Yes | 805 (17.0%) | 393 (16.8%) | 412 (17.2%) |  |
|  | NA | 355 | 120 | 235 |  |
| Ascites | No | 2983 (58.7%) | 1366 (55.5%) | 1617 (61.7%) | <10<sup>-3</sup> |
|  | Yes | 2098 (41.3%) | 1094 (44.5%) | 1004 (38.3%) |  |
|  | NA | 16 | 5 | 11 |  |
| Encephalopathy | No | 4741 (93.7%) | 2302 (94.0%) | 2439 (93.4%) | 0.4211 |
|  | Yes | 318 ( 6.3%) | 147 ( 6.0%) | 171 ( 6.6%) |  |
|  | NA | 38 | 16 | 22 |  |
| Etiology | HCV | 4317 (91.6%) | 1981 (91.5%) | 2336 (91.6%) | 0.224 |
|  | HBV | 154 ( 3.3%) | 80 ( 3.7%) | 74 ( 2.9%) |  |
|  | HCV-HBV coinfection | 40 ( 0.8%) | 20 ( 0.9%) | 20 ( 0.8%) |  |
|  | Non-viral etiology | 204 ( 4.3%) | 84 ( 3.9%) | 120 ( 4.7%) |  |
|  | NA | 382 | 300 | 82 |  |

**Table 1 (continued):** Temporal changes in patients' general, clinical and tumor characteristics at HCC diagnosis.
|  |  | Year of HCC diagnosis |  |  |  |
| --- | --- | --- | --- | --- | --- |
|  |  | Overall | 2005-2013 | 2014-2019 |  |
|  |  | N = 5,097 | N = 2465 | N= 2632 | P-value |
| Imaging system used | CT Scan | 4892 (96.0%) | 2362 (95.8%) | 2530 (96.1%) | <10<sup>-3</sup> |
|  | MRI | 109 ( 2.1%) | 57 ( 2.3%) | 52 ( 2.0%) |  |
|  | Both | 51 ( 1.0%) | 6 ( 0.2%) | 45 ( 1.7%) |  |
|  | NA | 45 ( 0.9%) | 40 ( 1.6%) | 5 ( 0.2%) |  |
| Hepatic focal lesion (HFL) number | 1 | 2244 (45.4%) | 1061 (44.4%) | 1183 (46.3%) | 0.2744 |
|  | 2 | 602 (12.2%) | 284 (11.9%) | 318 (12.4%) |  |
|  | 3 | 120 ( 2.4%) | 65 ( 2.7%) | 55 ( 2.2%) |  |
|  | >3 (multifocal) | 1979 (40.0%) | 978 (41.0%) | 1001 (39.1%) |  |
|  | NA | NA: 152 | 77 | 75 |  |
| Largest HFL (cm) | Median [Q25-Q75] | 5 [3-7.5] | 5.60 [3.50-8.00] | 5.00 [3.00-7.00] | <10<sup>-3</sup> |
|  | NA | 2472 | 1082 | 1390 |  |
| Vascular invasion | No | 3811 (75.9%) | 1834 (75.5%) | 1977 (76.3%) | 0.5094 |
|  | Yes | 1207 (24.1%) | 594 (24.5%) | 613 (23.7%) |  |
|  | NA | 79 | 37 | 42 |  |
| Distant metastasis | No | 4092 (81.9%) | 1960 (81.0%) | 2132 (82.8%) | 0.0918 |
|  | Yes | 902 (18.1%) | 460 (19.0%) | 442 (17.2%) |  |
|  | NA | 103 | 45 | 58 |  |
| BCLC stage at diagnosis | O/A | 848 (17.1%) | 366 (15.3%) | 482 (18.8%) | <10<sup>-3</sup> |
|  | B | 754 (15.2%) | 282 (11.8%) | 472 (18.4%) |  |
|  | C | 1478 (29.8%) | 625 (26.2%) | 853 (33.3%) |  |
|  | D | 1872 (37.8%) | 1114 (46.7%) | 758 (29.6%) |  |
|  | NA | 145 | 78 | 67 |  |

Changes in the distribution of BCLC stages at HCC diagnosis over time are presented in Figure 1 and Table 1. Late stage diagnosis decreased over time: BCLC stage D represented 46.7% of the patients before 2005-2013 vs. 29.6% in 2014-2019 (p<10^−3^).

Changes in laboratory values at HCC diagnosis are presented in Supplementary Figure S3. We observed significant decreases over time in alanine aminotransferase (ALT), asparate aminotransferase (AST), alpha-fetoprotein (AFP) and total bilirubin titers, while albumin, prothrombin, and platelets concentrations increased over time (all p<10^−3^).

Time changes in the treatment option by BCLC stage at diagnosis are displayed in Figure 2. Curative therapies were nearly exclusively decided among patients diagnosed with BCLC stages 0 to D, while patients diagnosed with BCLC stages C or D received almost exclusively noncurative locoregional therapies or supportive care, with no substantial change over time.

**Figure 2:**
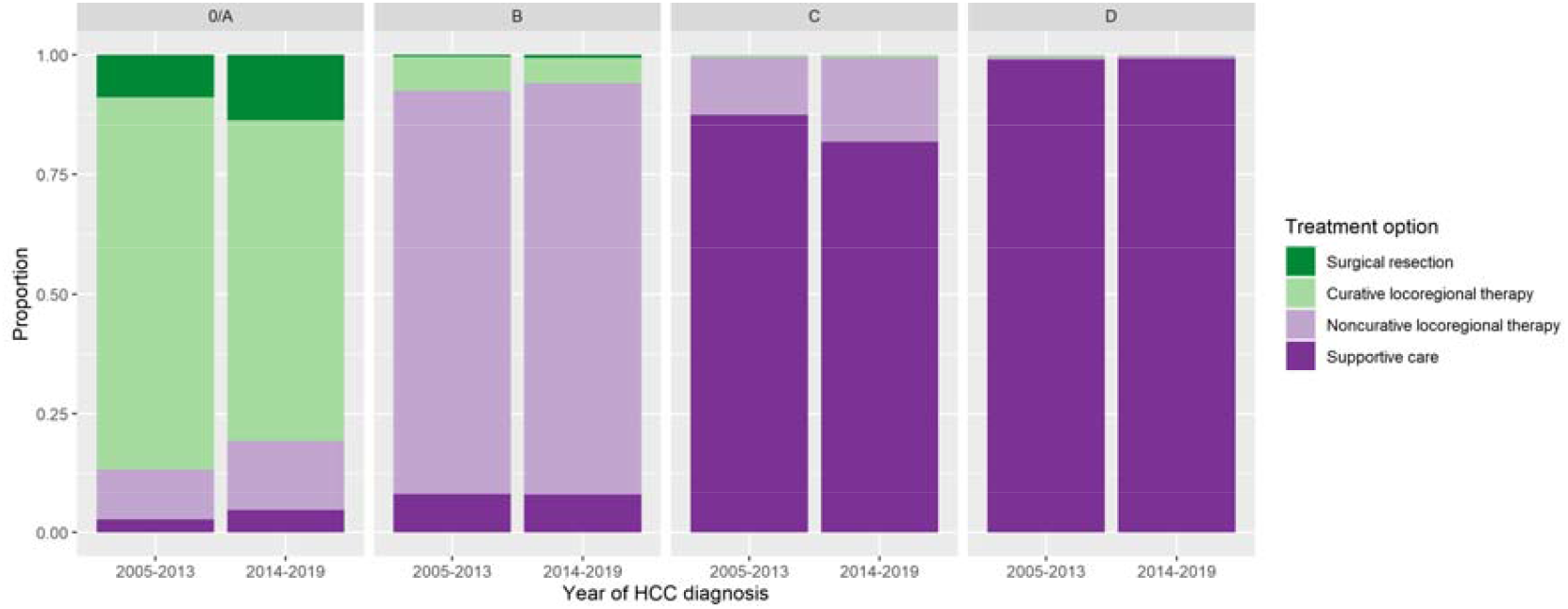
Distribution of treatment option by year of HCC diagnosis and BCLC stage at HCC diagnosis.

### Survival

A total of 4,651 deaths were observed among the 5,097 patients included in the survival analysis (91.2%). Overall, the median survival was 8.1 months (95% confidence interval, CI: 7.8-8.4) after HCC diagnosis. One year survival was 38% overall, and in patients with BCLC stages 0/A, B, C, D, it was 83%, 63%, 30% and 14%, respectively (Table 2). Kaplan-Meier survival curves are displayed in Figure 3, which shows that the proportional hazard assumption is not valid for the “*period of diagnosis*” variable (p<10^−3^), so the Cox regression model was not suitable. BCLC stage at diagnosis was significantly associated with overall survival, so was the type of treatment received (both p<10^−3^). Patients who received a curative treatment (either surgical ablation or curative locoregional therapy) experienced a median survival of 30.7 months (CI: 28.4-32.5). Figure 3 also shows that the period of diagnosis was not associated with overall survival (p=0.47), as one could have expected, considering the higher proportion of BCLC stages 0/A or B at diagnosis in the later vs. earlier period. When stratifying on BCLC stage at diagnosis, we observed a significantly higher survival in the 2005-2013 vs. 2014-2019 period among those diagnosed at BCLC stage 0/A (p<10^−3^) and B (p=0.029), but not for the stages C or D (Figure S4).

**Table 2:** Median and 6- to 24-months survival overall and by sub-groups.

|  | Median survival,<br>months (95% CI) | 6-month survival<br>(95% CI) | 12-month<br>survival (95% CI) | 24-month<br>survival (95% CI) |
| --- | --- | --- | --- | --- |
| **All** | 8.1 (7.8-8.4) | 59% (58-61) | 38% (37-39) | 18% (17-19) |
| **Year of HCC diagnosis** |  |  |  |  |
| 2005-2013 | 7.3 (6.9-7.8) | 56% (54-58) | 36% (34-38) | 19% (18-21) |
| 2014-2019 | 8.8 (8.4-9.3) | 62% (60-64) | 40% (38-42) | 17% (15-18) |
| **BCLC stage at diagnosis** |  |  |  |  |
| O/A | 28.0 (26.7-29.9) | 94% (93-96) | 83% (81-86) | 59% (56-63) |
| B | 15.3 (14.2-16.4) | 88% (85-90) | 63% (60-67) | 27% (24-30) |
| C | 7.2 ( 6.8- 7.8) | 57% (55-60) | 30% (27-32) | 9% (8-11) |
| D | 4.0 ( 3.6- 4.2) | 34% (32-36) | 14% (13-16) | 4% (3-5) |
| **Etiology** |  |  |  |  |
| HCV | 8.2 (7.8- 8.6) | 59% (58-61) | 38% (36-39) | 17% (16-19) |
| HBV | 6.0 (4.3- 8.3) | 49% (41-57) | 33% (26-41) | 19% (14-27) |
| HBV-HCV | 7.2 (4.6-10.2) | 55% (42-74) | 26% (15-45) | 12% (5-29) |
| Non-viral etiology | 6.2 (4.4- 8.4) | 51% (44-58) | 35% (29-43) | 15% (10-21) |
| **Treatment option** |  |  |  |  |
| Ablation | 31.4 (29.2-33.4) | 96% (94-97) | 87% (84-89) | 64% (60-68) |
| Surgical resection | 27.6 (22.1-40.0) | 91% (85-97) | 81% (73-90) | 59% (49-71) |
| TACE | 17.1 (15.8-17.8) | 89% (87-91) | 67% (64-70) | 30% (27-33) |
| Supportive care | 4.7 ( 4.6- 5.0) | 41% (39-43) | 18% (17-19) | 4% (4-5) |
| Multiple treatment | 11.1 ( 9.1-15.5) | 72% (65-79) | 47% (40-56) | 22% (16-30) |
| None/Other/Unknown | 7.2 ( 5.5- 8.0) | 55% (49-62) | 25% (20-32) | 9% (6-14) |

**Figure 3:**
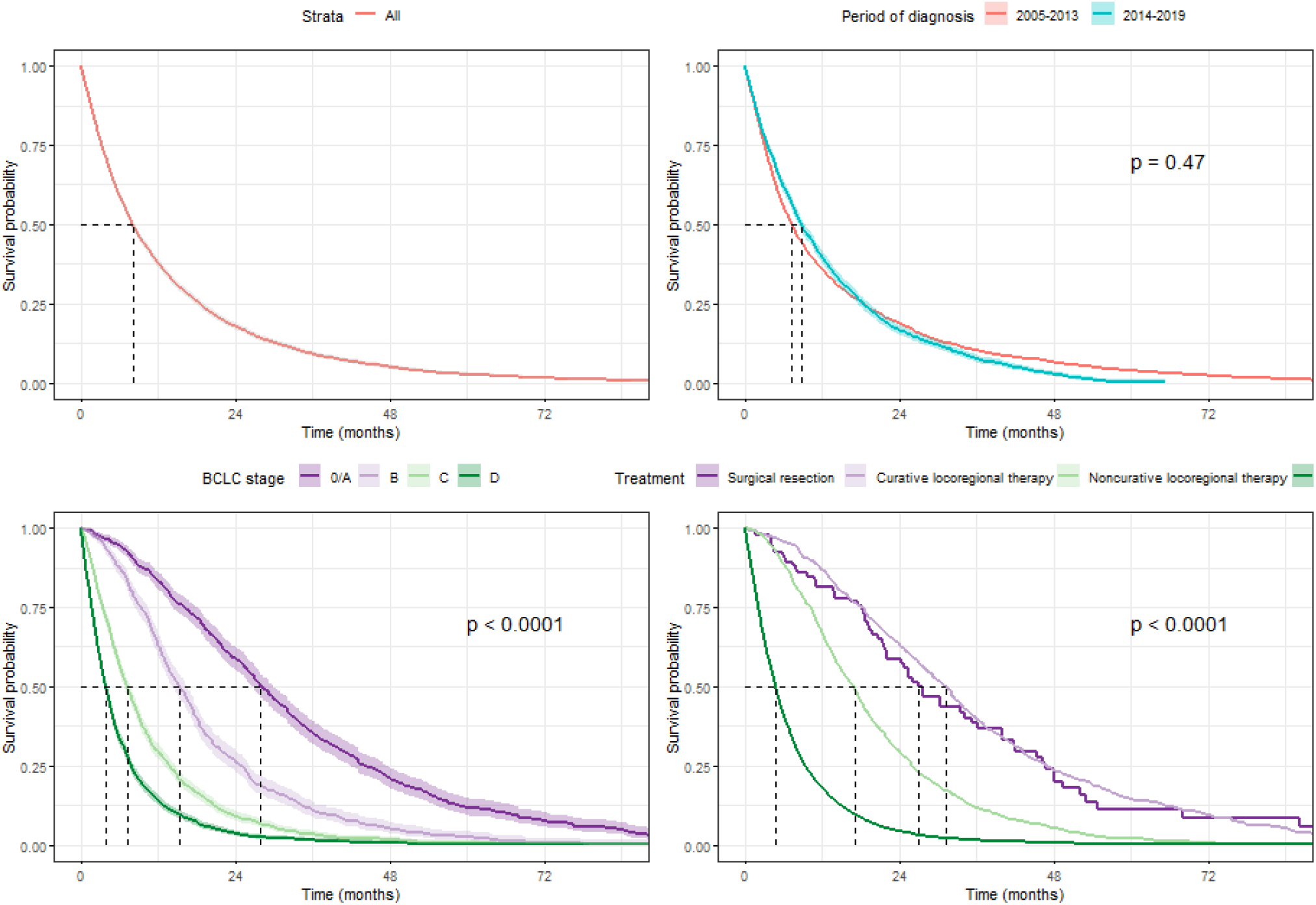
Kaplan-Meier survival curves: for the whole studied population (A), by period of diagnosis (B), by BCLC stage at HCC diagnosis (C), by treatment option (D). P-value computed using the log-rank test.

The differences in RMST according to individual and tumor characteristics are presented in Table 3. The multivariablebl analysis included 4,433 patients (87.0%) with complete information regarding the variables included in the model. Upon-admission factors independently associated with a lower mean survival time were age, tobacco consumption, vascular invasion and distant metastasis (all p<10^−3^). Patients with hypertension had a mean survival superior by 1.36 months (95% CI: 0.37; 2.34) to patients without. Diabetes was not associated with survival (p=0.79). HCC diagnosis in 2014-2019 was independently associated with a reduced survival time, with a life expectancy equivalent to 2.59 (95% CI: 1.37; 3.81) months shorter than patients diagnosed in 2005-2013. We observed a strong gradient associating advanced BCLC stage at diagnosis with reduced survival. For instance, those diagnosed at BCLC stage D had a mean survival inferior by 19.23 months (95% CI: -20.50; -17.96) compared to those diagnosed at BCLC stage A. In the stratified analysis, all these associations were consistently observed before and after 2014 (Supplementary Table 1).

**Table 3 :** Association between mortality and general, clinical and tumor characteristics. RMST : Restricted mean survival time.

|  |  | Crude RMST | p | Adjusted RMST | p |
| --- | --- | --- | --- | --- | --- |
| **Sex** | Men | ref. |  | ref. |  |
| | Women | 2.54 ( 1.09; 3.99) | $<10^{-3}$ | -0.33 ( -1.23; 0.57) | 0.4699 |
| **Area of residence** | Urban | ref. |  | ref. |  |
|  | Rural | -1.04 ( -2.42; 0.35) | 0.1413 | -0.36 ( -1.15; 0.44) | 0.3803 |
| **Age** | (per 10y. increase) | -1.96 ( -2.41; -1.51) | $<10^{-3}$ | -0.97 ( -1.37; -0.57) | $<10^{-3}$ |
| **Period of HCC diagnosis** | 2005-2013 | ref. |  | ref. |  |
| | 2014-2019 | -0.04 ( -0.78; 0.71) | 0.9257 | -2.30 ( -3.02; -1.57) | $<10^{-3}$ |
| **Tobacco consumption** | No | ref. |  | ref. |  |
| | Yes | -3.96 ( -5.02; -2.90) | $<10^{-3}$ | -1.62 ( -2.37; -0.87) | $<10^{-3}$ |
| **Hypertension** | No | ref. |  | ref. |  |
| | Yes | 2.92 ( 1.36; 4.47) | $<10^{-3}$ | 1.51 ( 0.60; 2.42) | 0.0012 |
| **Diabetes** | No | ref. |  | ref. |  |
|  | Yes | 2.18 ( 0.77; 3.58) | 0.0025 | 0.11 ( -0.69; 0.92) | 0.7868 |
| **Etiology** | HCV | ref. |  | ref. |  |
|  | HBV | -0.71 ( -3.79; 2.38) | 0.6534 | -0.18 ( -2.19; 1.84) | 0.8628 |
|  | HBV-HCV | -1.06 ( -7.40; 5.28) | 0.7442 | 0.99 ( -2.67; 4.65) | 0.5968 |
|  | non-viral | -1.59 ( -4.32; 1.15) | 0.2552 | -1.08 ( -2.65; 0.50) | 0.1794 |
| **Vascular invasion** | No | ref. |  | ref. |  |
| | Yes | -7.64 ( -8.59; -6.68) | $<10^{-3}$ | -2.28 ( -2.97; -1.59) | $<10^{-3}$ |
| **Distant metastasis** | No | ref. |  | ref. |  |
| | Yes | -7.90 ( -8.83; -6.96) | $<10^{-3}$ | -2.03 ( -2.75; -1.31) | $<10^{-3}$ |
| **BCLC stage at diagnosis** | O/A | ref. |  | ref. |  |
| | B | -7.78 ( -9.51; -6.04) | $<10^{-3}$ | -8.60 ( -10.10; -7.10) | $<10^{-3}$ |
| | C | -14.52 ( -16.18; -12.86) | $<10^{-3}$ | -15.77 ( -17.06; -14.48) | $<10^{-3}$ |
| | D | -19.25 ( -20.79; -17.72) | $<10^{-3}$ | -19.23 ( -20.50; -17.96) | $<10^{-3}$ |

### HCC recurrence free survival

Among 819 patients (16.1% of the sample) who received a curative therapy (surgical resection: 13.2%; curative locoregional therapy: 86.8%), HCC recurrence was recorded among 451 (55.1%). The median recurrence-free survival was 22.1 months (95% CI: 20.1-25.6) and was not significantly different among those with an initial HCC diagnosis in 2014-2019 vs. 2005-2013 (p=0.12) (Figure 4).

**Figure 4:**
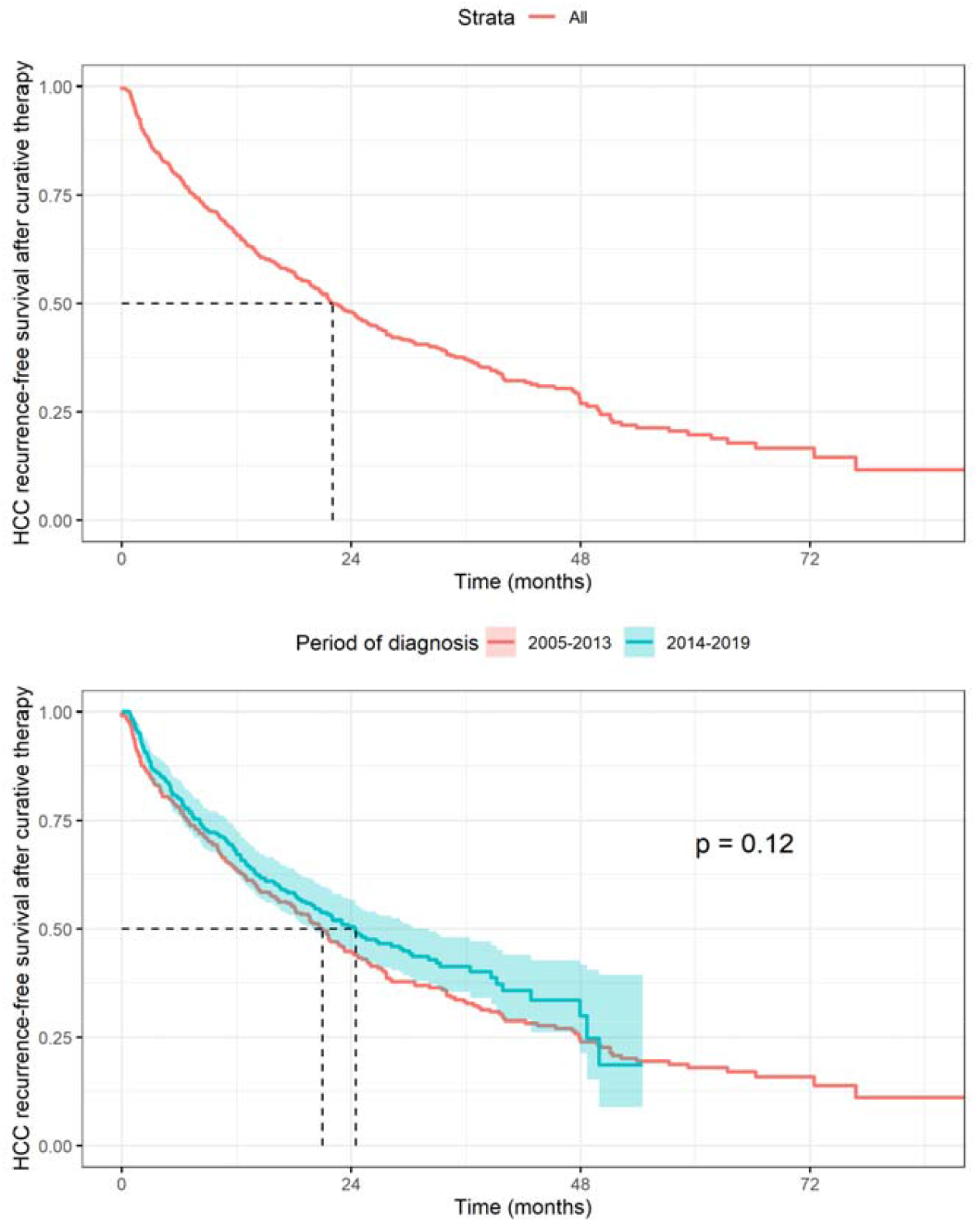
Kaplan-Meier curves for HCC recurrence-free survival after curative therapy, overall (top) and by period of diagnosis (bottom). P-value computed using the log-rank test

## DISCUSSION

In this monocentric, retrospective study, we assessed changes in presentation and survival of patients diagnosed with HCC between 2005 and 2019 in a region with a high disease incidence. We observed significant trends over time in individual characteristics at diagnosis, such as older age, a higher proportion of urban patients, a lower proportion of smokers, and higher proportions of metabolic disorders (diabetes, hypertension) in the recent period. Moreover, we observed an important trend toward HCC diagnosed at earlier stages in the recent period. Overall, the patient’s survival after the HCC diagnosis was poor, with a median of 8.1 months. The BCLC stage at diagnosis constituted a performant predictor of survival in this patient’s population. Despite a trend toward HCC diagnosed at earlier stages, we did not observe a significant improvement of survival over time, which constitutes a challenging observation. Overall, treatments offered in this medical center were in line with international guidelines, and 16.1% of the patients who received a curative treatment presented an improved survival. However, HCC recurrence was frequent among patients cured for HCC, with a median time to recurrence of 22 months.

Several temporal trends and consistencies have been observed in the presentation of a patient diagnosed with HCC during the period covered by the study. The etiology of HCC remained largely driven by HCV (>90%). This predominant etiology and the history of the Egyptian HCV epidemic linked to anti-schistosomal control activities in the 1960s–70s also explain the shift toward older age at HCC diagnosis. Trends toward more frequent metabolic disorders, mainly diabetes and hypertension, is likely linked to global trends in overweight, one of their main risk factors, which has been reported in Northern Africa as in most regions in the world over the past decades [23]. One of our main observations within this cohort is the significant trend toward earlier diagnosed HCC during the recent period (2014-2019). This trend has to be related to the 2014 initiation of the national DAA scale-up program, which resulted in a dramatic increase of DAA-treated patients from 45,000 in 2014 to 577,000 in 2016 [24]. Within the Egyptian HCV model of care, treated cirrhotic patients are regularly monitored for detection of complications of cirrhosis or HCC, which may have contributed to such a trend toward earlier diagnosis [14]. However, even in the recent period, nearly one third of the patients were diagnosed at a terminal stage of the disease (BCLC stage D), with no treatment option beyond best supportive care. This highlights the remaining efforts for HCC surveillance.

Overall, in this retrospective cohort, we document an 8.1-months median survival after HCC diagnosis, which appears lower than previously reported in Egyptian studies. Gomaa et al. documented a median survival of 15 months in 2010-2012 [12], and Waziry et al estimated the median survival following HCC at 22.8 months in 2013-2014 [24]. However, both studies were conducted at the National Liver Institute in Menoufiya, one of the largest centers for the treatment of HCV infection, among which patients were diagnosed at earlier BCLC stage as compared to our study (BCLC stage 0/A: 25% and 56%, respectively as compared to the 17% reported here).

In our study sample, the BCLC staging system performed remarkably well in estimating patient prognosis. Indeed, the BCLC stage at diagnostic was the most discriminating independent factor predicting survival. It exhibited a strong gradient, ranging from a median survival of 28 months for BCLC stages 0/A to 4 months for stage D. However, as compared to those reported in high-income countries, the BCLC stage-specific survival we document here was slightly lower (for instance: 7 months for BCLC stage C here vs. >1 year reported in [16]).

The multivariableble survival analysis revealed several independent prognostic factors after HCC diagnosis. Tobacco consumption has long been identified as a risk factor for HCC incidence, but its independent association with mortality after HCC diagnosis has not been consistently reported in previous study, probably due to small sample sizes [12,14,25]. The strong statistical power allowed by our large sample size may explain that we could report an independent association between tobacco consumption and mortality after HCC diagnosis, even if the effect size is small (mean survival reduced by 1.6 months) and only opens modest perspectives for secondary prevention. We did not observe an independent association between diabetes and the evolution of HCC, in line with previous evidence [9]. However, we observed a significant, though modest, better prognostic among patients with baseline hypertension (mean survival improved by 1.5 months), contradicting the results observed European patients [9]. The biological plausibility of hypertension as a favorable prognostic factor for survival after HCC diagnosis remains questionable though, and might be better explicated by a differential, potentially improved, follow-up of patients with hypertension. In line with previous evidence, we did not report any difference in the prognostic based on the etiology of HCC [16,26].

Despite a trend toward earlier diagnosis of HCC in our population study, we did not observe an overall improvement in survival over time. Indeed, independently of the prognostic and risk factors discussed above, the later period of diagnosis (2014-2019) was associated with a mean survival reduced by 2.3 months compared to the earlier period. Our analysis stratified by BCLC stage at diagnosis revealed that this poorer survival in the recent period was driven by a poorer survival among patients diagnosed at BCLC stage 0/A (and more modestly by patients diagnosed at stage B), while survival among patients at advanced or terminal stage of the disease remained comparable across periods (Supplementary figure 4). This results suggest that the distribution of one or several risk factor for survival after HCC diagnoses, which were not collected in our study, may have changed over time.

We acknowledge several limitations to these results. Principally, the monocentric nature of our study may limit the generalisability of our results beyond the catchment population of the Damietta referral center. We relied on retrospective data collection based on medical file, for which the degree of completion varied over time. For instance, etiology of HCC was more frequently unknown in the pre- vs. post-2014 period. However, we relied on criteria ensuring specific eligibility of patients over the study period, and final inclusion rate in the analysis was high (97.8%), reducing the risk of a possible inclusion bias.

Moreover, in the time-to-event analysis, the event was documented among >90% of patients, thus limiting the risk of bias due to informative censoring [27]. Furthermore, with more than 5,000 inclusions, this is the most extensive study conducted in Egypt to estimate the survival of patients following HCC diagnosis. We measured correlates of mortality using RMST difference, which interpretation is more intuitive than hazard ratios. It is increasingly recognized that hazard ratios from Cox proportional hazards models represent a potential threat to causal inference, even in the absence of unmeasured confounding, measurement error, and model misspecification [28].HCC remains one of the significant causes of malignancy in Egypt. The perspectives of novel promising treatment strategies are limited [16], and timely diagnosis is key to ensuring the best treatment strategy under the available options. The present study documents the progress toward earlier HCC diagnoses made in Egypt since 2014 and the opportunity that the DAA strategy has generated. In the meantime, with more than half of the patients diagnosed at an advance or terminal stage of the disease in the latest years, the current study also weights the remaining efforts to ensure the optimal benefits of HCC care and treatment.

## Supporting information

STROBE checklist

## Data Availability

All data produced in the present study are available upon reasonable request to the authors.

## Ethical clearance

This study received approval from the Research Ethics Committee for Human Subject Research at Air Force Specialized Hospital and from the Egyptian Ministry of Health on 31st Jul 2019 and on October, 9th 2019 respectively.

## Author contribution

KJ, AT, LBLN, AF and MEK designed the study. AF obtained funding; provided clinical management to patients and collected the patient data. AT and MEK ensured fieldwork coordination. KJ and LT ensured data curation. KJ performed the statistical analysis. KJ drafted the manuscript. All authors approved the final version of this article, including the authorship list.

## Data availability

Data related to this study is available upon a reasonable request through an email to the corresponding author

## SUPPLEMENTARY MATERIAL

**Supplementary Figure 1:**
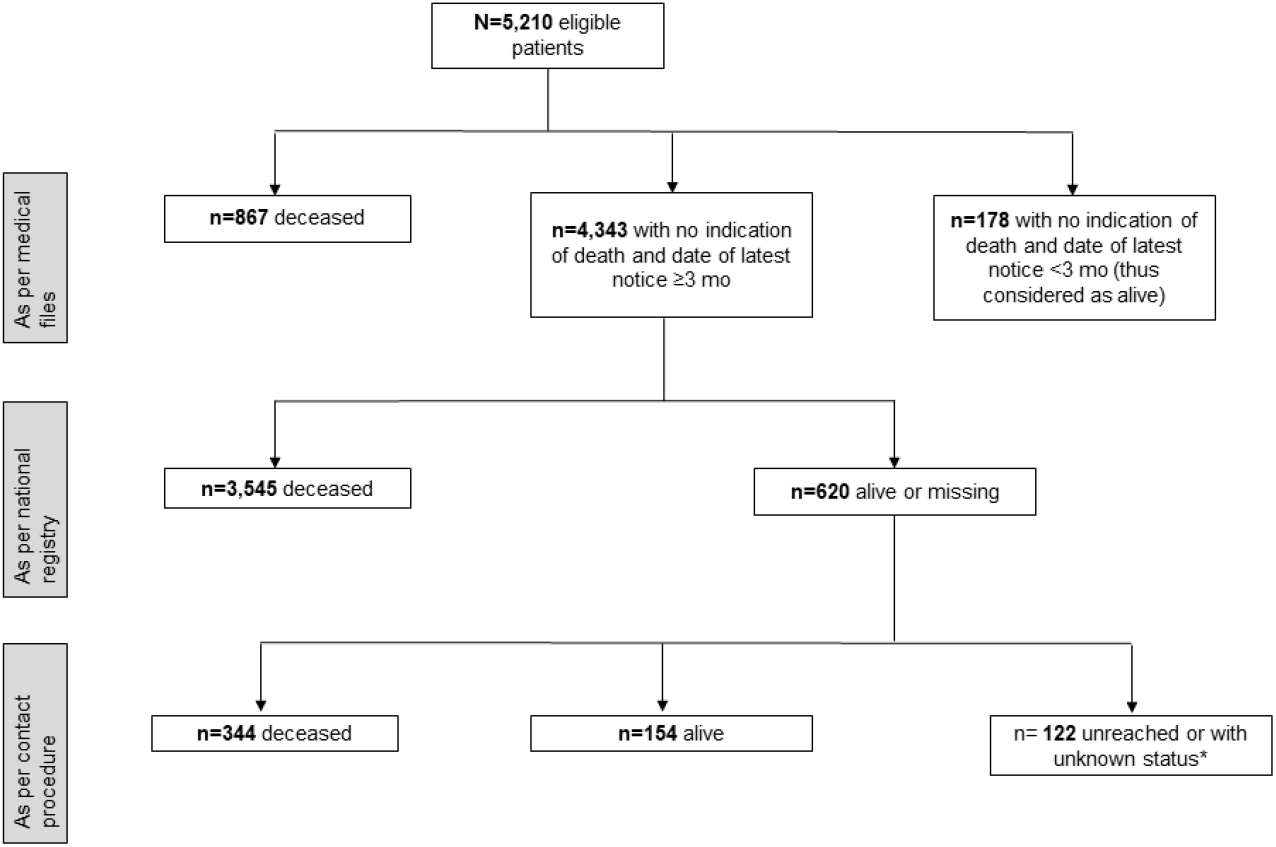
Flowchart of vital status ascertainment.

**Supplementary Figure 2:**
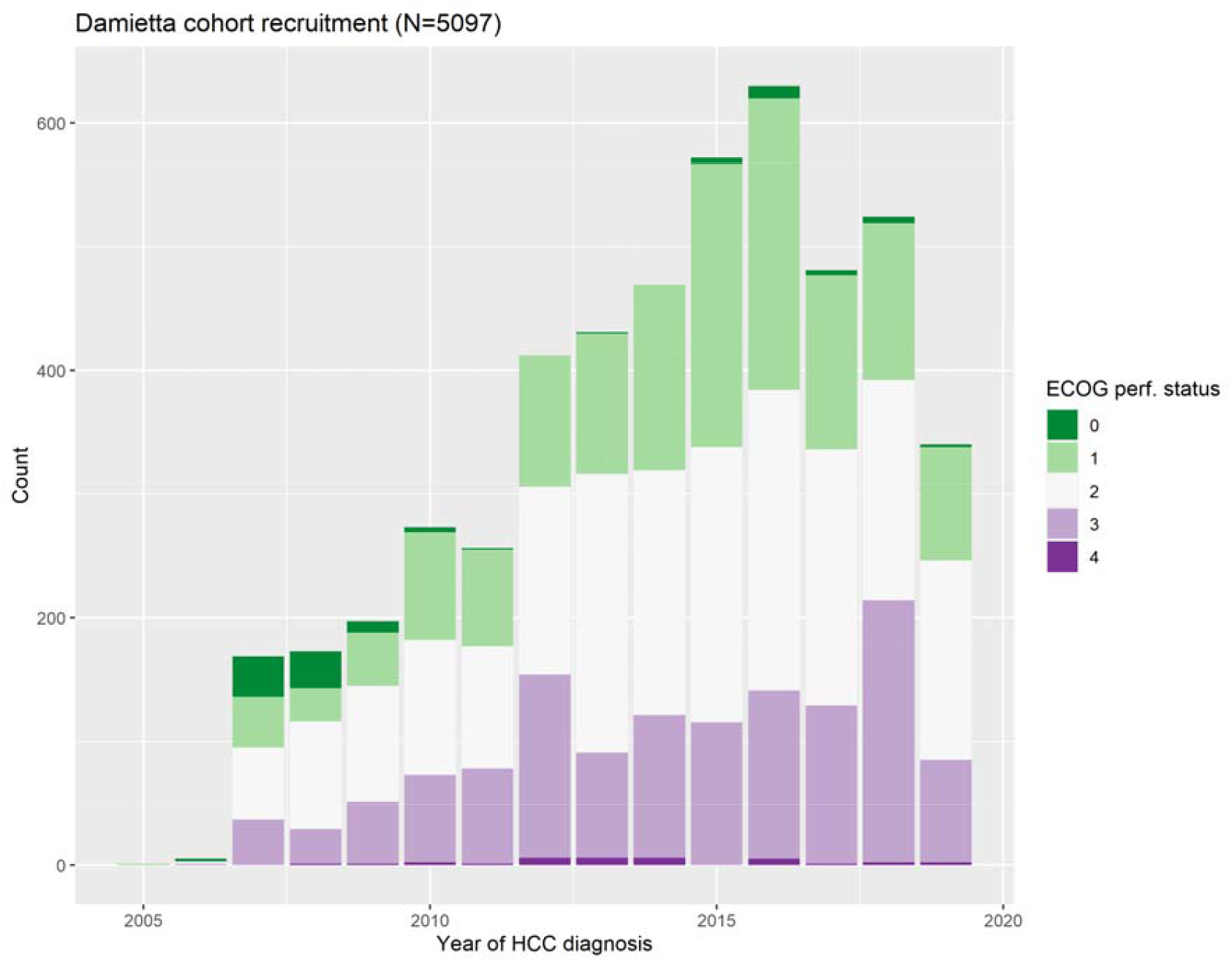
Distribution of ECOG performance status at HCC diagnosis, by year of diagnosis.

**Supplementary figure 3:**
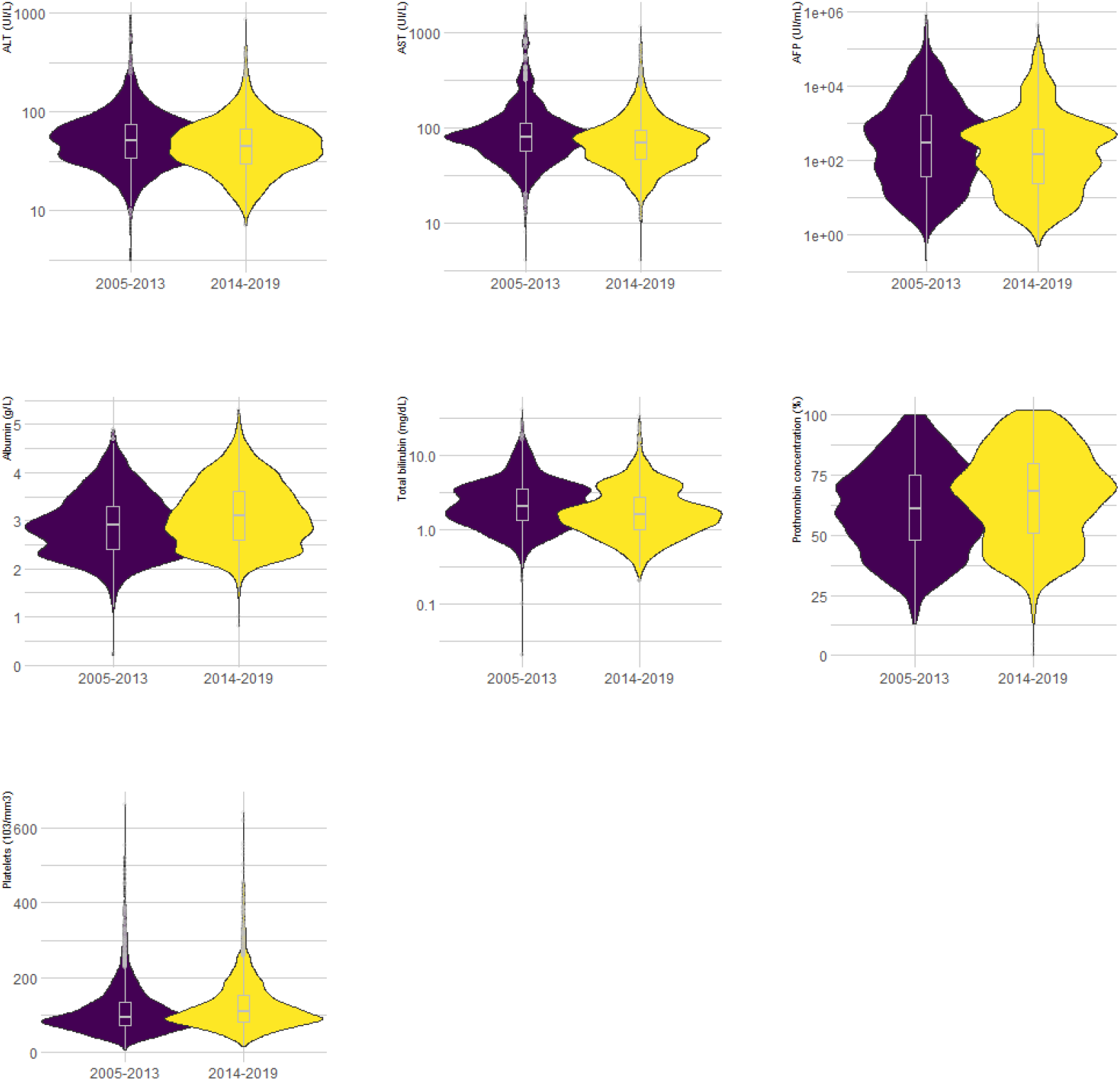
Temporal changes in laboratory values at time of HCC diagnosis (all Student test p-values <10^−3^).

**Supplementary Figure 4:**
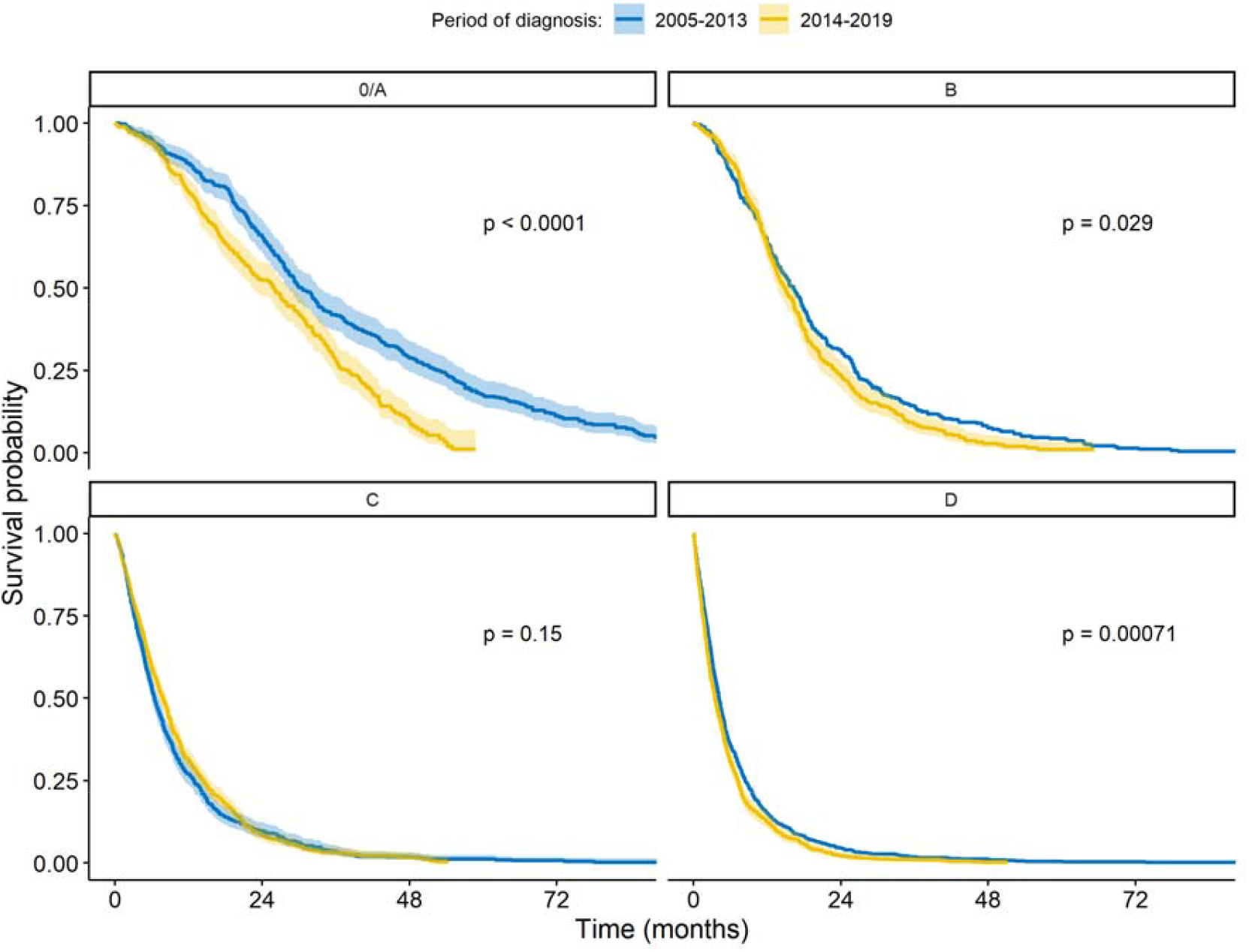
Survival per time period and BCLC stage at HCC diagnosis.

**Supplementary Table 1:**
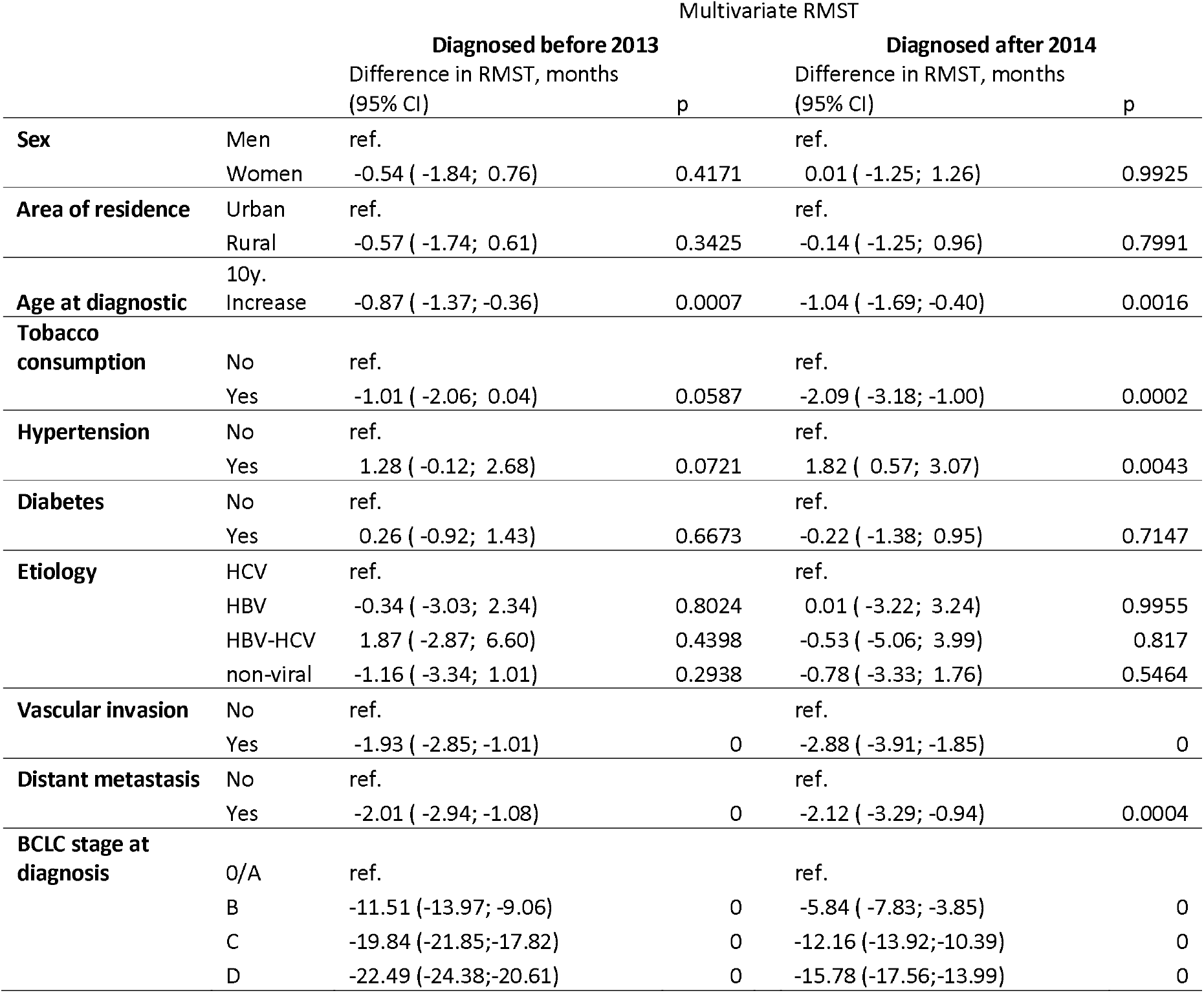
Association between mortality and general, clinical and tumor characteristics, stratified on the period of HCC diagnosis.

